# Tumor-Origin.com: A Machine Learning Platform for Predicting Tumor Tissue of Origin from Somatic Mutation Profiles

**DOI:** 10.64898/2025.12.27.25343092

**Authors:** Saicharan Vellanki, Peter Feiszt, Paraic A. Kenny

## Abstract

Standard pathology workup sometimes fails to definitively identify tumor tissue-of-origin in cancers with ambiguous diagnoses or unknown primary sites, complicating treatment decisions. Molecular assays can aid diagnosis but require additional tissue and increase healthcare costs. Intending to leverage routinely collected somatic mutation profiles from comprehensive genomic profiling, we developed Tumor-Origin.com, a machine learning platform to predict tumor tissue-of-origin from mutation data alone. We trained five classifiers on 10,945 tumor mutation profiles from the MSK-IMPACT cohort and validated performance on an independent set of 770 tumors from the Gundersen Precision Oncology cohort spanning 52 cancer types. Performance was strongest for the most common tumor types, reflecting their relative over-representation in training data. Among cancer types with more than five cases, the Logistic Regression classifier achieved the highest average top-3 accuracy of 49%, followed by the Support Vector Machine at 43%. At least one algorithm delivered ≥40% accuracy in 23 cancer types. Our integrated platform thus provides robust tumor origin predictions across diverse cancers. We have implemented a web-based tool (https://tumor-origin.com) to assist clinicians and researchers in refining diagnoses of cancers of unknown primary without requiring additional tissue or costly testing.

## INTRODUCTION

Approximately 3-5% of new cancer cases are “cancers of unknown primary”, which is associated with treatment uncertainty and typically poor outcomes (Kato et al., 2021; Raghav, 2025). A variety of approaches are used to clarify the diagnostic uncertainty, with the goal of offering patients a more disease-tailored treatment regimen, rather than empiric chemotherapy. Sequential immunohistochemistry testing with panels of antibodies evaluating various differentiation markers has long been a diagnostic mainstay in this situation (Lin and Liu, 2014). More recently, a variety of molecular diagnostics have been used, including gene expression profiling, epigenomics, and various integrations thereof (Kato et al., 2021). Like most advanced cancer cases in the modern era, these tumors are also frequently subjected to next-generation sequencing with the goal of identifying genetic biomarkers predictive of therapeutic benefit (Chakravarty and Solit, 2021).

Each of these approaches requires consumption of additional biopsy specimens and/or ordering of additional clinical testing, leading to concerns about both tissue stewardship (Kerr et al., 2024) and expense (Abrams et al., 2021; Ehman et al., 2024). Here we describe a straightforward tool which works with NGS data that is now typically already captured during routine care. We trained five machine learning algorithms on tumor mutation profiles of 10,945 cancer cases. Entry of the list of mutated genes reported on a comprehensive genome profiling report results in a quantitative prediction from each algorithm regarding the likelihood that the mutation profile corresponds to tumors from a variety of organs. We validated the approach using a series of 770 tumors from the Gundersen Precision Oncology cohort (Feiszt and Kenny, 2025). A web version of the application is available at http://tumor-origin.com.

## MATERIALS & METHODS

### Training Data

The algorithms were trained on genomic data from the MSK-IMPACT clinical sequencing cohort (Zehir et al., 2017) consisting of 10,945 tumor specimens from 10,336 patients. Data were downloaded from https://www.cbioportal.org/study/summary?id=msk_impact_2017. Data were preprocessed to generate a binarized matrix for 338 genes in which 0 indicated no reported alteration and 1 indicated an alteration of that gene in the tumor.

### Algorithms

Five supervised machine learning classifiers—K-Nearest Neighbors (KNN), Decision Tree, Logistic Regression, Support Vector Machine (SVM) with calibrated probabilities, and a OneVsRestClassifier wrapping a Decision Tree—were trained and deployed as an integrated AI platform for tumor origin prediction. Processed mutation vectors are fed into each of these trained models, which independently generate probability scores for all possible tumor origins. All models aggregate their own outputs to produce a ranked list of the top three most likely primary sites. Performance evaluation was conducted using the top-3 accuracy metric.

Software: All code was written in Python and is available from https://github.com/paraickenny/TumorOriginPredictor

### Ethical Approval

The Gundersen Precision Oncology Cohort has been approved by the Institutional Review Board of the Gundersen Health System (PI: Kenny, IRB # 2-22-02-003).

## RESULTS

### Algorithm training

This MSK-IMPACT dataset contained mutational profiles of 338 genes from 10,945 tumor samples spanning 27 different cancer types. The dataset was converted into a binary mutation matrix, with 1 indicating a gene mutation and 0 indicating a non-mutated gene. All reported alterations (single nucleotide variants, indels, amplifications, and deletions) were all scored as 1. This binarized matrix served as the input feature set for training. Using Python, five supervised machine learning classifiers—K-Nearest Neighbors, Decision Tree, Logistic Regression, Support Vector Machine with calibrated probabilities, and a OneVsRestClassifier wrapping a Decision Tree—were trained on this data. The training process involved fitting each model to the binary mutation profiles with corresponding tumor tissue labels. After training, the models were serialized and saved using Python’s pickle module to enable efficient loading and prediction without retraining, ensuring reproducibility and stability for future use.

### Validation

A validation dataset was also curated, containing 770 de-identified patient samples across 52 unique tissue origins, coming from the Gundersen Precision Oncology cohort. This cohort consists of patients who had tumor next-generation sequencing performed as part of their routine cancer care. Data from clinical reports were abstracted and data for each gene in each case was binarized (mutant/non-mutant) as described above. To facilitate rapid processing of these data, some additional code (batch_analysis.py in the github repository) was added to enable batch processing and tabulation of the resulting output.

There were some differences in the ways tissue of origin was categorized in both datasets, which required harmonization to enable a robust comparison. For example, lung tumors were categorized as small cell, non-small cell, and mesothelioma in the MSK dataset and were all grouped as “Lung Cancer” in the Gundersen dataset. Head and neck tumors were grouped as a single group in the MSK dataset, but subdivided into distinct anatomical locations in the Gundersen dataset. We pre-specified a cross-walk matrix for acceptable tissue matches prior to computing the results (Supplemental Table 1).

The performance of each algorithm in the test cohort is shown in Figure 1. In general, performance was strongest in the cases of the most common tumor types in this cohort, which likely reflects a similar over-representation of these cancer types in the training cohort. Among cancer types with more than 5 cases in the validation cohort, the Logistic Regression classifier performed best, with an average accuracy of 49% as judged by inclusion of the correct answer among the top three predictions, followed by the Support Vector Machine classifier at 43%. At least one algorithm delivered at least 40% accuracy in 23 cancer types in the cohort.

**Figure 1.**
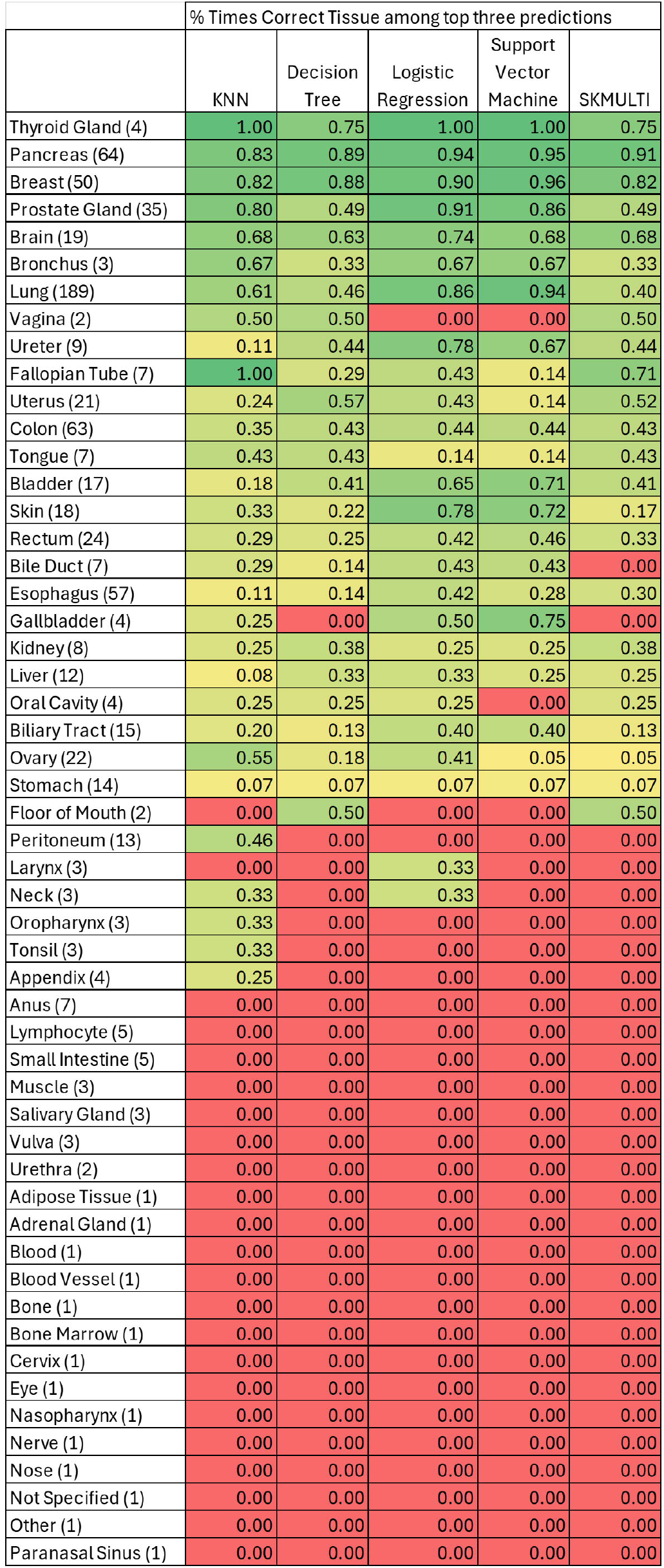
Evaluation of prediction accuracy using a validation dataset from the Gundersen Precision Oncology Cohort. Mutations from 770 Gundersen Precision Oncology Cohort tumors were evaluated using each of the five algorithms trained on MSK-IMPACT data. The percentage of cases where the correct tissue of origin was among the top three predictions is shown, ranked by the median value for each tumor type among all algorithms. The number of cases of each cancer type included in the cohort is indicated in the left column.

### Deployment of web-based prediction tool

We developed a public-facing implementation of this approach and hosted it on Microsoft Azure, enabling clinicians and researchers to use the platform to evaluate cancers of unknown primary and ambiguous metastatic tumors. Users simply enter patient mutation data, and they receive tumor origin predictions within seconds (Figure 2). In addition, all rank-ordered probabilities are output, enabling direct review of probability predictions in cases where one or more candidate tissues of origin are already included in the differential diagnosis. The platform can be accessed at https://tumor-origin.com.

**Figure 2.**
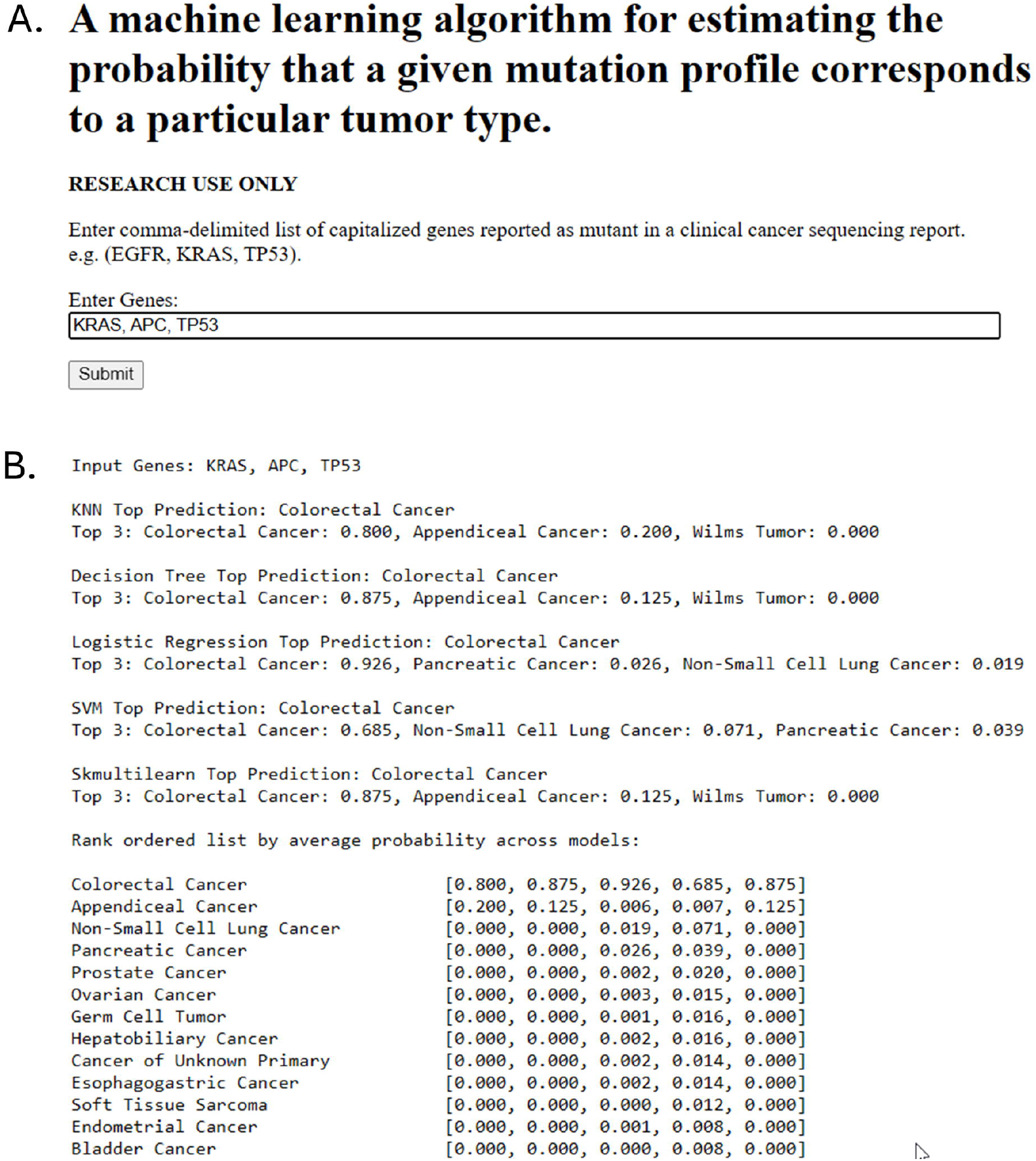
The tumor-origin.com web browser interface. (A) Data entry screen showing evaluation of a tumor with KRAS, APC, and TP53 mutations. (B) Results output showing the top prediction for each model, the top 3 predictions for each model, and the probabilities assigned to lower ranked candidate tissues-of-origin.

## DISCUSSION

We demonstrate the utility of this straightforward approach of using classifiers trained on a large, well-curated tumor genomics dataset to evaluate candidate tissues-of-origin for cancers of unknown primary. Validation in the independent Gundersen Precision Oncology Cohort showed that this approach performs well, especially for common tumor types that are well-represented in the training dataset. This approach should be considered complementary to other diagnostic tools developed for this purpose, including immunohistochemical analysis with antibody panels, RNA-sequencing, and epigenetic analysis (Kato et al., 2021). Notably, our approach uses data typically already collected during NGS analysis of cancers of unknown primary, and does not require the utilization of additional biopsy specimens or expensive molecular testing. Our web-based implementation enables users to evaluate this approach using their own data.

One limitation is that the preparation of the binarized matrix for training relied on the somatic mutation calls from the MSK_IMPACT data, some of which may have included variants of uncertain significance. A more rigorous attempt to fully exclude VUS might be expected to at least modestly improve algorithm performance. Variants of Uncertain Significance were not included in the Gundersen Precision Oncology Cohort dataset.

A further limitation was the difference in homogeneity between the training (single platform) and test (a real world cohort tested on multiple platforms with different numbers of genes from vendors including STRATA Oncoogy, Tempus and Foundation Medicine) cohorts. The inclusion of cases in the validation cohort which had been tested on NGS panels smaller than the MSK-IMPACT panel almost certainly resulted in some missing data, where one of the MSK-IMPACT genes was mutated but not tested in the sample. This limitation should bias the validation toward poorer performance, so we believe that the performance estimates provided herein probably somewhat underestimate the likely performance achievable in datasets developed with consistently larger panels.

In summary, Tumor-Origin.com is an effective tool that uses routinely collected somatic mutation data and machine learning models trained on a large dataset to predict tumor tissue-of-origin. Its web-based platform complements existing diagnostics without needing extra tissue or costly tests, providing easy access for clinicians and researchers to support personalized cancer care.

## Data Availability

All data produced in the present study are available upon reasonable request to the authors.

## ACKNOWLEDGEMENTS

This study was funded by the Gundersen Medical Foundation. PK holds the Dr. Jon & Betty Kabara Endowed Chair in Precision Oncology.

## Notes

### Competing Interest Statement

The authors have declared no competing interest.

### Author Declarations

The Institutional Review Board of Gundersen Clinic, Ltd. gave ethical approval for this work.

